# Pre-infusion Exhaled breath volatile organic compounds predict severe CRS and ICANS after CAR T-cell therapy

**DOI:** 10.64898/2026.05.28.26354352

**Authors:** Amalia Z. Berna, Johannes F. Fahrmann, Ehsan Irajizad, Hamid Khoshfekr Rudsari, Yang Liu, Joe Logan, Khaled Murtada, Jonathan Grandy, Matthew Edwards, Amy Ayers, Sairah Ahmed, Sattva S. Neelapu, Neeraj Y. Saini, Audrey Odom John, Teny M. John

**Affiliations:** Division of Infectious Diseases, Children’s Hospital of Philadelphia, Philadelphia, Pennsylvania, USA; Department of Clinical Cancer Prevention, The University of Texas MD Anderson Cancer Center, Houston, Texas, USA; Department of Biostatistics, The University of Texas MD Anderson Cancer Center, Houston, Texas, USA; SepSolve Analytical Ltd, 1060 Guelph Street, Kitchener, Ontario, N2B 2E3, Canada; Department of Lymphoma/Myeloma, The University of Texas MD Anderson Cancer Center, Houston, Texas, USA; Division of Infectious Diseases, Department of Pediatrics, Perelman School of Medicine, University of Pennsylvania, Philadelphia, Pennsylvania, USA; Department of Infectious Diseases, Infection Control and Employee Health, The University of Texas MD Anderson Cancer Center, Houston, Texas, USA

## Abstract

**Background:** Severe cytokine release syndrome (CRS) and immune effector cell–associated neurotoxicity syndrome (ICANS) are major dose-limiting toxicities of chimeric antigen receptor (CAR) T-cell therapy. Existing pre-infusion biomarkers offer modest discrimination, motivating non-invasive alternatives.

**Methods:** We prospectively enrolled 26 patients with relapsed/refractory large B-cell lymphoma receiving axicabtagene ciloleucel. Pre-infusion (day −1) exhaled breath samples were analyzed by gas chromatography–mass spectrometry for 40 volatile organic compounds (VOCs). Candidates with univariate AUC > 0.65 for severe (grade ≥2) CRS or ICANS were carried forward to sensitivity-maximization-at-given-specificity with LASSO regularization (SMAGS-LASSO), which selected separate panels for each outcome. Model performance was assessed by leave-one-out cross-validation with permutation p-values and Harrell bootstrap optimism correction.

**Results:** The 4-VOC CRS panel (heptanal, benzaldehyde, 2-butanone, ethylbenzene) achieved LOOCV AUC 82.5% (80% sensitivity at 88% specificity) and the 3-VOC ICANS panel (nonanal, allyl methyl sulfide, levomenthol) achieved AUC 86.3% (67% sensitivity at 86% specificity). By tertile, severe CRS occurred in 8/9 (89%) high-risk versus 2/9 (22%) low-risk patients (Cox HR 6.82, 95% CI 1.41–32.9, p=0.017) and severe ICANS occurred in 8/9 (89%) versus 2/9 (22%) (HR 8.28, 95% CI 1.73–39.6, p=0.008). Each 1-SD score increase corresponded to a 3.80-fold higher hazard of severe CRS (p<0.001) and 4.36-fold higher hazard of severe ICANS (p<0.001). In head-to-head comparison, the 3-VOC ICANS panel outperformed the modified Endothelial Activation and Stress Index (mEASIX) (ΔAUC +0.36, DeLong 1-sided p=0.008). The 4-VOC CRS panel had numerically higher AUC than mEASIX (ΔAUC +0.19, p=0.150).

**Conclusions:** Pre-infusion exhaled breath VOC panels stratify CAR T-cell recipients by severity and timing of severe CRS and ICANS, providing a non-invasive complement to existing serum biomarkers. Multi-institutional validation is warranted.

**Key Points:**

1. Pre-infusion breath VOC panels predict severe CRS (AUC 82.5%) and ICANS (AUC 86.3%).
2. Day −1 panel scores predict not only severity but also faster onset of severe toxicity, identifying high-risk patients before infusion.

## Background

Chimeric antigen receptor (CAR) T-cell therapy has transformed the treatment of relapsed or refractory lymphoid malignancies^1^. Its broader adoption is limited by immune-mediated toxicities, namely cytokine release syndrome (CRS) and immune effector cell–associated neurotoxicity syndrome (ICANS), that cause substantial morbidity and mortality.^1,2^

Early risk stratification is necessary because the window for preemptive intervention is narrow. CRS typically begins in the first week after CAR T-cell infusion, with median onset of 3 days,^3^ and ICANS commonly follows between day 4 and day 7. Untreated low-grade CRS can progress to grade 3 or 4 disease and ICANS, increasing the risk of intensive care admission, prolonged hospitalization, and death ^2^. Diagnosis of CRS and ICANS is currently clinical based on symptoms and signs, and severity is graded according to ASTCT criteria^4^. For both syndromes, grade ≥2 disease is defined as severe and clinically significant that requires specific interventions including tocilizumab for CRS and corticosteroids for ICANS ^5^. Existing laboratory biomarkers are insufficient for clinical decision-making: C-reactive protein rises in essentially all patients with CRS but lags clinical changes by at least 12 hours, and cytokine assays (interleukin [IL]-6, IL-8, IL-10, interferon-γ, monocyte chemoattractant protein-1) are available only in specialized laboratories^5,6^. The modified endothelial activation and stress index (mEASIX) calculated as lactate dehydrogenase x C-reactive protein / platelet count) at pre-infusion lymphodepletion has been proposed as a composite laboratory predictor of CAR T-cell toxicity^7^. But population-based external validation demonstrated substantially lower performance, with area under the curves (AUCs) of 0.61 – 0.62 for ICANS and no significant CRS risk stratification^8^. As a single pre-infusion snapshot of endothelial activation derived entirely from routine laboratory values, mEASIX cannot capture the dynamic metabolic changes before and after CAR T-cell expansion, highlighting the need for novel biomarker approaches capable of individualized, real-time risk stratification.

Exhaled breath contains thousands of volatile organic compounds (VOCs) generated by cellular metabolism. Lung-origin VOCs are released directly in to the airways, while systemically produced VOCs reach the alveolar gas via diffusion across the alveolar-capillary barrier before exhalation^9^. Reproducible alterations in breath VOCs have been described in patients with CRS secondary to COVID-19^10^, which shares clinical features with CAR T-cell–induced CRS^11^. Breath analysis provide a rapid, non-invasive method to predict CRS and ICANS before CAR T-cell infusion, when prophylactic intervention is still possible. In this proof-of-concept study, we evaluated the utility of exhaled breath VOCs measured before CAR T-cell infusion for predicting clinically significant (grade ≥ 2) CRS and ICANS in patients with lymphoid malignancies receiving axicabtagene ciloleucel (axi-cel) and established a predictive model from breath profiles by thermal desorption–mass spectrometry.

## Methods

### Study population

We prospectively enrolled patients with relapsed or refractory large B-cell lymphoma receiving axicabtagene ciloleucel (axi-cel) at MD Anderson Cancer Center from March 2023 to June 2024. Exclusion criteria were an active second cancer of a different type and inability to provide a breath sample because of altered mental status. Breath samples were collected from 29 enrolled patients; 3 samples failed baseline quality control (excess sample humidity), resulting in 26 patients with analyzable breath samples in the analytic cohort. Breath samples were collected according to established protocols at two timepoints relative to CAR T-cell infusion: day −1 (post-lymphodepletion, and before infusion) and day +1 (per protocol, with a 48-hour collection window) (Figure 1). Day −1 samples were used to derive the predictive 4-VOC CRS panel and 3-VOC ICANS panel using sensitivity-maximization-at-given-specificity with LASSO regularization (SMAGS-LASSO) under leave-one-out cross-validation; day +1 samples were used to characterize how breath VOC biology changes during the peri-infusion window and were not used for model training or validation. All participants provided informed consent, and the study was approved by the MD Anderson Cancer Center Institutional Review Board (IRB 2022-0801).

**Figure 1.**
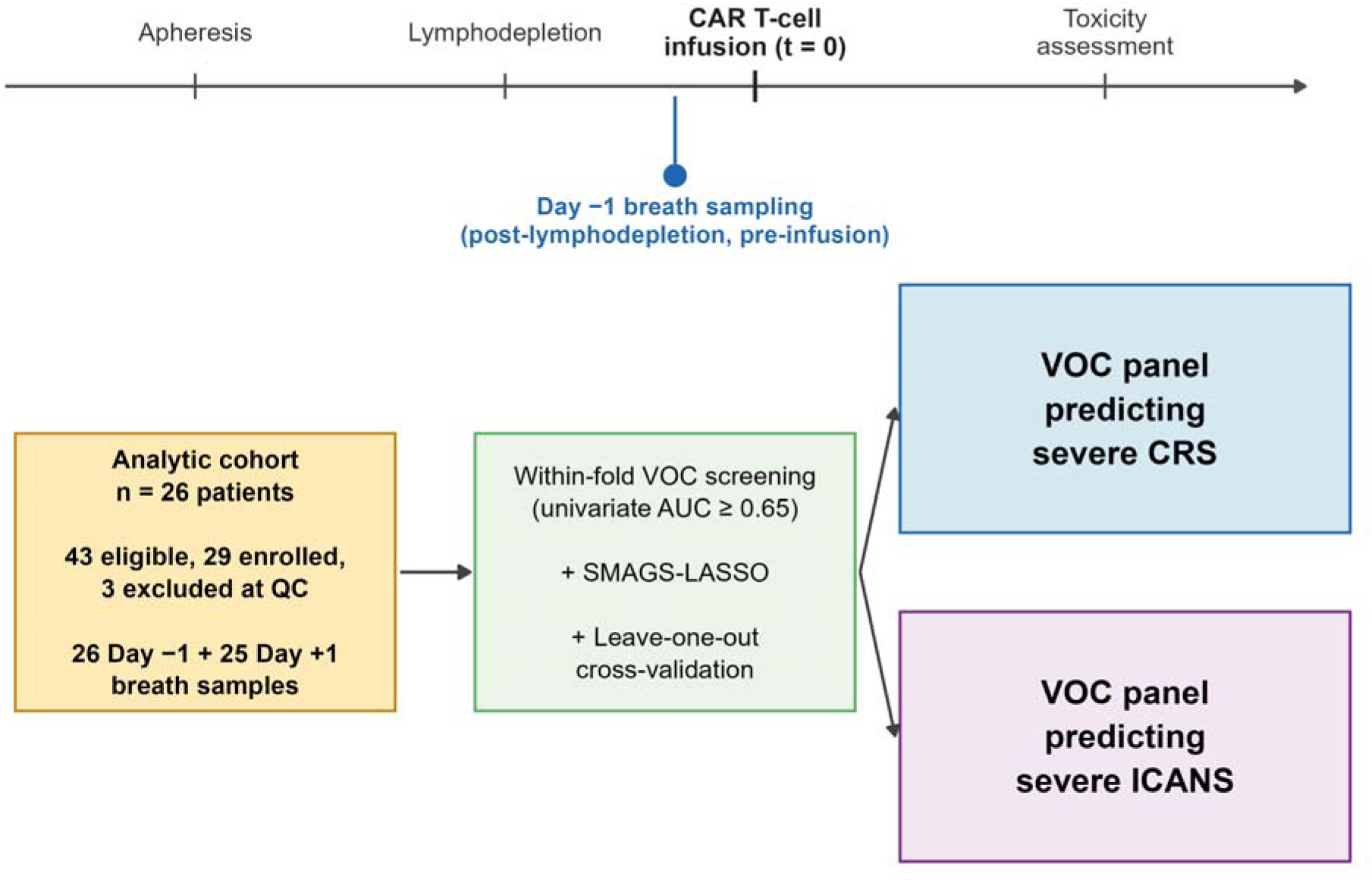
Study schema: Breath was collected on day −1 post-lymphodepletion, pre-infusion and VOCs were quantified by TD-GC/MS. Day−1 VOC profiles were used to train SMAGS-LASSO models predicting severe CRS and ICANS, with leave-out cross-validation.

### Toxicity grading

CRS and ICANS were graded according to the American Society for Transplantation and Cellular Therapy (ASTCT) consensus criteria^4^ and adjudicated by an institutional committee of stem cell transplant experts. Grade ≥2 was classified as severe because it prompts active intervention, including tocilizumab, corticosteroids, or intensive care monitoring).

### Sample collection

We collected breath and ambient air samples as previously described^12^. Briefly, patients actively exhaled into a 3-L SamplePro Flexfilm sample bag (SKC) and one liter of breath was discharged into three-bed universal sorbent tubes containing Tenax, Carbograph, and Carboxen (Markes International, UK). Ambient air was collected from the same room at the time of breath collection. Patients completed a 24-hour dietary recall to account for diet-derived VOCs.

### Thermal desorption and chromatography

Sorbent tubes were equilibrated to room temperature and loaded into an automated thermal desorption system (Ultra-xr, Markes International, UK). A gaseous internal standard mixture containing bromochloromethane, 1,4-difluorobenzene, chlorobenzene-d5, and 4-bromofluorobenzene was added to each tube immediately before analysis. Tubes were pre-purged with nitrogen for 10 min at 50 mL·min□¹ to minimize water interference. Thermal desorption was performed at 270 °C for 10 min, with analytes focused onto a Universal cold trap held at 10 °C and subsequently heated to 300 °C.

Chromatographic separation was performed by comprehensive two-dimensional gas chromatography (GC×GC) with a flow modulator, coupled to both a flame ionization detector and a time-of-flight mass spectrometer (BenchTOF, SepSolve, UK). The first-dimension column was a Stabilwax (30 m × 250 μm ID × 0.25 μm df); the second-dimension column was an Rtx-200 MS (5 m × 250 μm ID × 0.1 μm df). The oven was programmed from 40 °C to 215 °C at 3 °C·min□¹, then ramped to 260 °C; total runtime was 70 min. Helium carrier gas flow was 1.2 mL·min□¹ with a 2-second modulation period. Mass spectra were acquired by electron ionization at 70 eV, m/z 35–350, 50 Hz. Chemical standards were analyzed under identical conditions, and instrument performance was verified before each batch using external calibration standards.

### Breath analysis

Consistent with recent breath volatilome studies in clinical cohorts^13^, we adopted a targeted analytical approach, focused on a curated set of VOCs detectable in human breath, rather than an untargeted feature discovery, a pragmatic approach given the small cohort signal-to-noise considerations. Chromatographic data were analyzed using ChromSpace software (SepSolve Analytical Ltd, UK). A targeted semi-quantitative approach was applied to estimate compound concentrations (ppm). For each analyte, the peak area was normalized to that of a selected internal standard and multiplied by the known concentration of the internal standard to obtain the final concentration. Internal-standard assignments for all VOCs were harmonized across samples prior to analysis.

### Statistical analysis

We built two pre-infusion (day-1) VOC predictive panels, one for grade ≥ 2 CRS and one for grade ≥ 2 ICANS using sensitivity maximization at a given specificity with LASSO regularization (SMAGS-LASSO).^14,15^ The specificity constraint was set at ≥90% to relax specificity in favor of sensitivity, the clinically prioritized metric for early CAR T-cell toxicity prediction in which missing a high-risk patient (false negative) carries greater clinical cost than over-flagging a low-risk patient (false positive). L1 regularization (λ = 1.0) was applied to enforce feature sparsity, and the optimal coefficient set was selected from 216 optimizer combinations (9 methods × 8 tolerances × 3 Jacobian approximations) implemented in Python 3.10.

Within each leave-one-out cross-validation (LOOCV) fold, we first screened candidate VOCs by univariate AUC ≥ 0.65 in the training fold (to prevent test-fold leakage), then fit SMAGS-LASSO to the screened VOCs. Model performance was reported as the LOOCV AUC and permutation p-value (1,000 random label permutations). Model stability was assessed by Harrell’s bootstrap optimism correction with 1,000 resampling iterations. In each iteration, patients were sampled with replacement to form the training set, and patients not drawn in to the resample served as the held-out test set. We report the optimism-corrected AUC and the 95% bootstrap confidence interval (CI) of out-of-bag AUC (Supplementary Figure 9); 95% bootstrap CIs for sensitivity at the prespecified specificity were derived from 2,000 resamples.

To assess clinical risk stratification, we divided patients into low-, intermediate-, and high-risk tertiles defined by the 33rd and 67th percentiles of the day −1 panel score. Time from CAR T-cell infusion to grade ≥2 CRS or grade ≥2 ICANS was modeled by Cox proportional hazards regression with the day −1 panel score as the predictor (continuous per 1-SD increment, and as score tertiles), with non-events censored at day 14 (CRS) or day 28 (ICANS). Cumulative incidence by score tertile was estimated using Kaplan–Meier curves, and tertiles were compared by the log-rank test.

To characterize peri-infusion VOC biology, we compared day+1 breath samples with their day─1 counterparts using paired Wilcoxon signed-rank tests on raw concentrations and per-VOC univariate AUCs.

## Results

Patient characteristics are summarized in Table 1. Twenty-five of 26 patients (96%) developed any-grade CRS; 10 (38%) developed grade ≥2 CRS (median time to onset 4 days; range 1–7). Seventeen of 26 (65%) developed any-grade ICANS; 12 (46%) developed grade ≥2 ICANS (median time to onset 6 days; range 5–8). The timing of CRS and ICANS onset relative to breath sample collection is shown in Supplementary Figure 1.

**Table 1:**
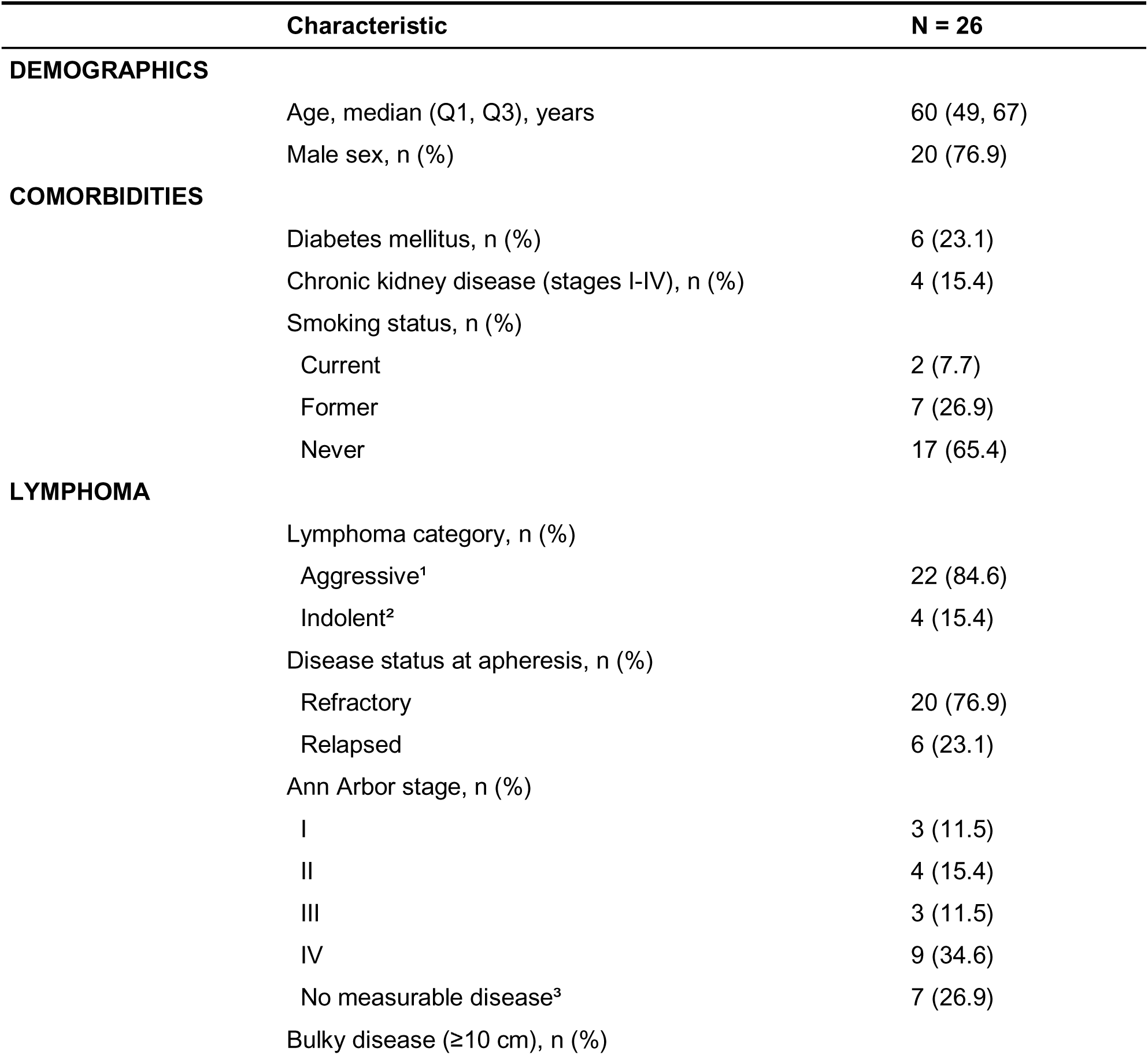
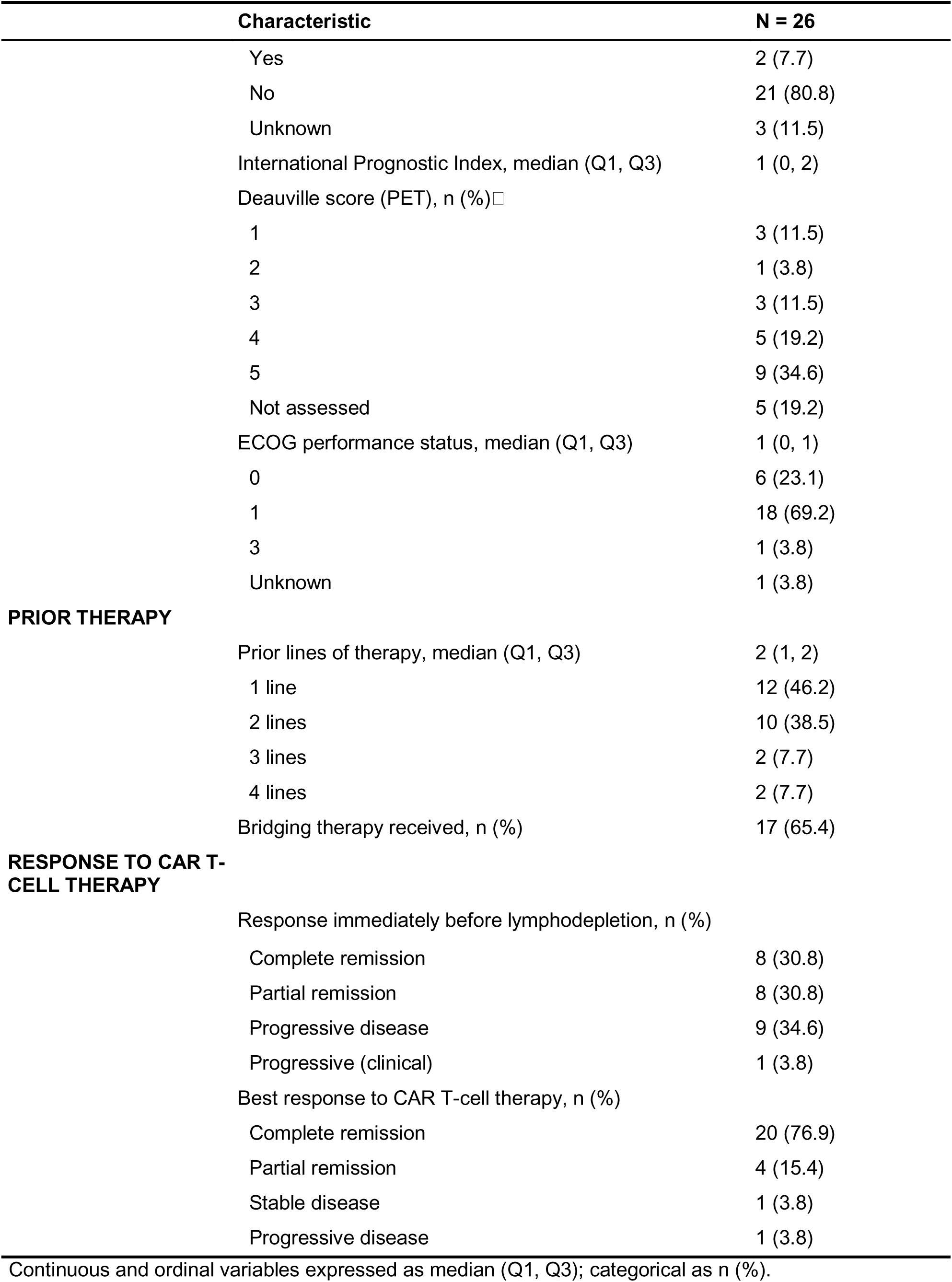
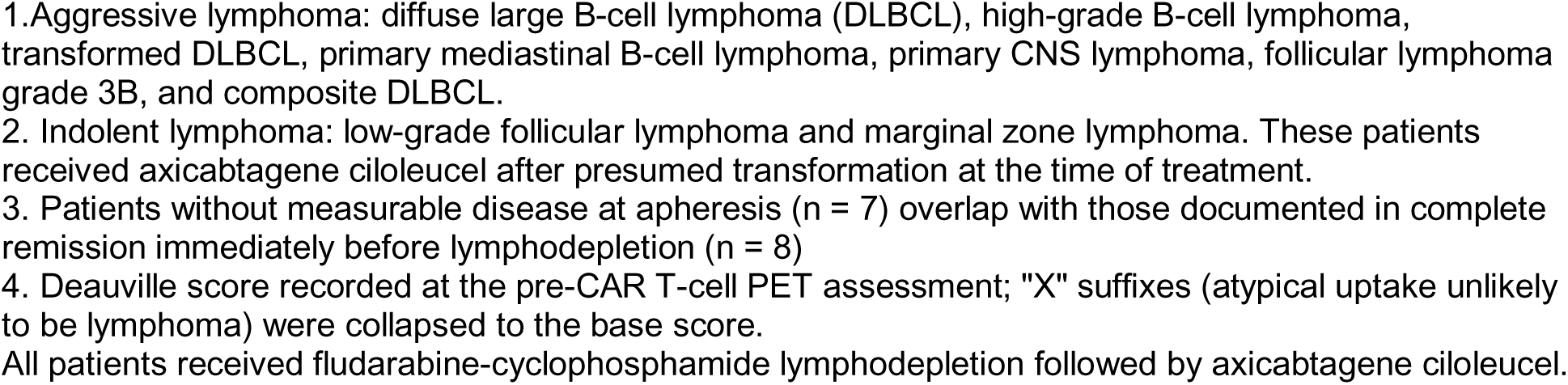
Baseline demographics of the cohort and response to CAR T-cell therapy.

### Day −1 breath VOCs predict grade ≥2 CRS

We compared day −1 breath profiles between patients who developed grade ≥2 CRS (n=10) and those who did not (n=16). Of the 40 VOCs included in the targeted breath analysis, 8 VOCs had univariate AUC ≥0.65, of which only heptanal reached nominal significance (Wilcoxon p=0.029). SMAGS-LASSO selected a 4-VOC panel (heptanal, benzaldehyde, 2-butanone, and ethylbenzene) with a LOOCV AUC of 82.5% (sensitivity 80% at ≥90% specificity, 95% bootstrap CI 50%–100%; permutation p=0.007; Harrell bootstrap-corrected AUC 0.877, 95% CI 0.73–0.96; Figure 2A, Supplementary Figure 9A). Day −1 panel score stratified patients into low-, intermediate-, and high-risk tertiles (defined by 33rd and 67th percentiles of the score distribution): grade ≥2 CRS occurred in 8 of 9 patients (89%) in the high-risk tertile, 0 of 8 (0%) in the intermediate-risk tertile, and 2 of 9 (22%) in the low-risk tertile (high- vs. low-risk tertile Cox HR = 6.82, 95% CI 1.41–32.9, p= 0.017; log-rank p = 2 × 10⁻□; Figure 2B–C). Higher scores were associated with faster onset of grade ≥ 2 CRS; by day 5, 56% (5/9) had developed severe CRS, compared with 0% in the intermediate-risk and 11% (1/9) in the low-risk tertile (Cox HR 3.80, 95% CI 1.76–8.20, p = 7 × 10⁻□).

**Figure 2.**
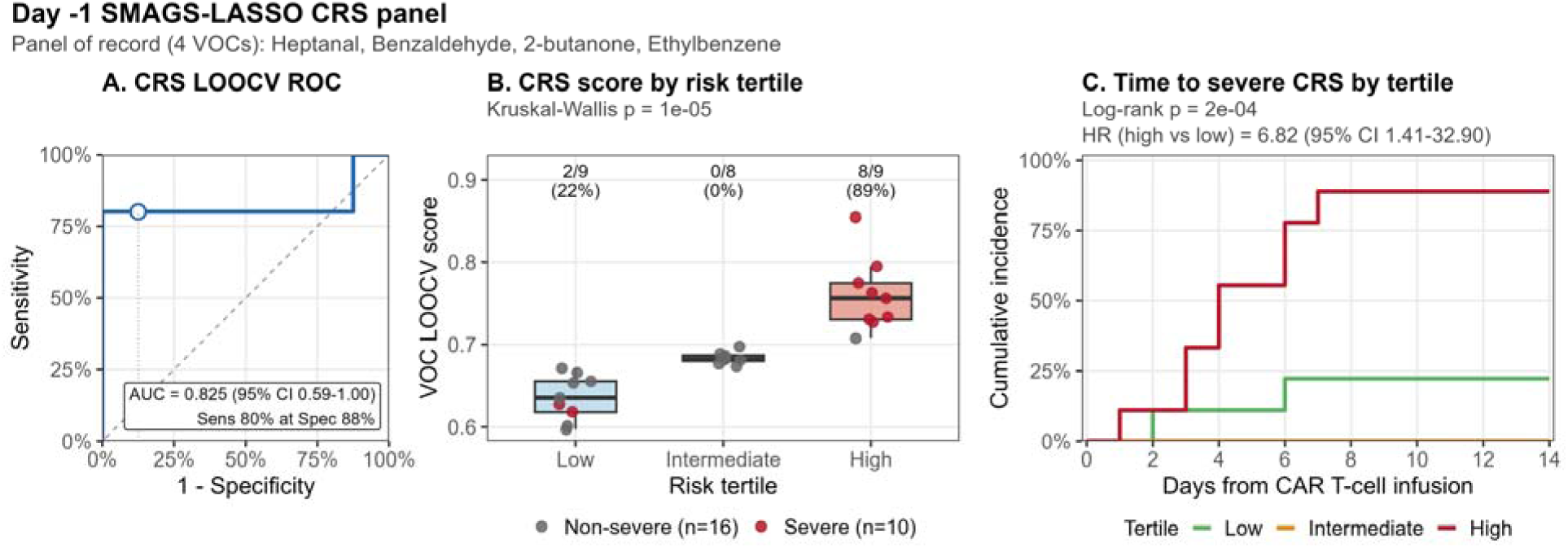
Day −1 SMAGS-LASSO CRS panel performance (N = 26; 10 severe-CRS events). Pre-infusion (Day −1) breath VOC panel for predicting grade ≥2 CRS, comprising four compounds (heptanal, benzaldehyde, 2-butanone, and ethylbenzene) selected by within-fold univariate screening (AUC ≥ 0.65) followed by SMAGS-LASSO model fitting. (A) Leave-one-out cross-validated receiver-operating-characteristic curve; LOOCV AUC and 95% DeLong confidence interval are shown inset. (B) Distribution of LOOCV-derived panel scores by predicted-risk tertile, with patients colored by severe-CRS status; the proportion of patients with grade ≥2 CRS in each tertile is annotated above the box, and the Kruskal–Wallis test for score differences across tertiles is shown in the subtitle. (C) Kaplan–Meier cumulative-incidence curves for time to grade ≥2 CRS by panel-score tertile, with patients censored at day 14. The log-rank p-value and the Cox proportional-hazards hazard ratio (high vs low tertile, 95% confidence interval) are shown inset. CRS, cytokine release syndrome; LOOCV, leave-one-out cross-validation; AUC, area under the receiver-operating-characteristic curve.

### Day −1 breath VOCs predict grade ≥2 ICANS

Of the 40 VOCs in the targeted breath analysis, 12 had univariate AUC ≥0.65 for ICANS prediction. SMAGS-LASSO selected a 3-VOC ICANS panel (nonanal, allyl methyl sulfide, and levomenthol) with a LOOCV AUC of 86.3% (sensitivity 67% at ≥90% specificity, 95% bootstrap CI 44%–100%; 26/26 folds; permutation p<0.001; Harrell bootstrap-corrected AUC 0.867, 95% CI 0.74–0.91; Figure 3A, Supplementary Figure 9B).

**Figure 3.**
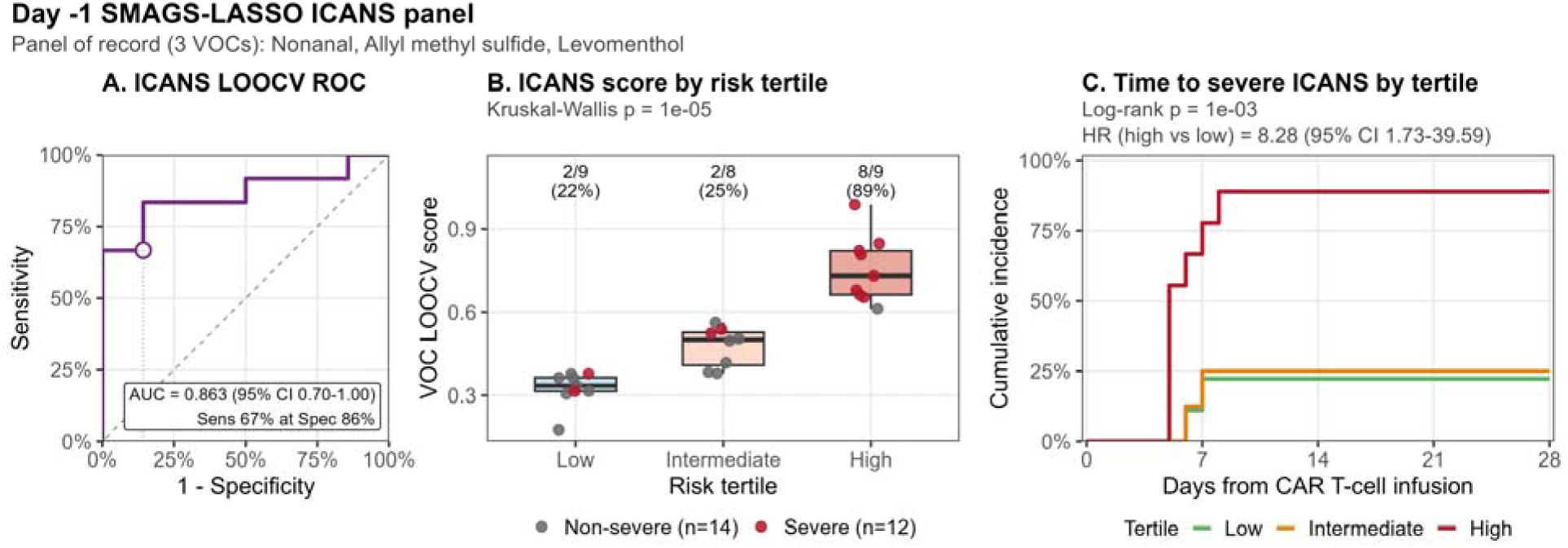
Day −1 SMAGS-LASSO ICANS panel performance (N = 26; 12 severe-ICANS events). Pre-infusion (day −1) breath VOC panel for predicting grade ≥2 ICANS, comprising three compounds, nonanal, allyl methyl sulfide, and levomenthol, selected by within-fold univariate screening (AUC ≥ 0.65) followed by SMAGS-LASSO model fitting. (A) Leave-one-out cross-validated receiver-operating-characteristic curve; LOOCV AUC and 95% DeLong confidence interval are shown inset. (B) Distribution of LOOCV-derived panel scores by predicted-risk tertile, with patients colored by severe-ICANS status; the proportion of patients with grade ≥2 ICANS in each tertile is annotated above the box, and the Kruskal–Wallis test for score differences across tertiles is shown in the subtitle. (C) Kaplan–Meier cumulative-incidence curves for time to grade ≥2 ICANS by panel-score tertile, with patients censored at day 28. The log-rank p-value and the Cox proportional-hazards hazard ratio (high vs low tertile, 95% confidence interval) are shown inset.

As with CRS, day −1 panel scores similarly stratified patients by severe ICANS risk; grade ≥ 2 ICANS occurred in 8 of 9 patients (89%) in the high-risk tertile, 2 of 8 (25%) in the intermediate-risk tertile, and 2 of 9 (22%) in the low-risk tertile (high- vs. low- tertile Cox HR = 8.28, 95% CI 1.73–39.6, p = 0.008; log-rank p = 0.001; Figure 3B,C). Onset of severe ICANS was earlier in the high-risk tertile; by day 5, 56% (5/9) had developed severe ICANS, while no patients in the intermediate-risk or low-risk tertiles had (Cox HR per 1-SD = 4.36, 95% CI 2.09–9.13, p < 0.001; Figure 3C). The ICANS panel shared no compounds with the CRS panel, indicating that severe grade ≥2 CRS and severe grade ≥2 ICANS arise from distinct pre-infusion metabolic states.

### Peri-infusion VOC biology evolves between day −1 and day +1

In the 25 patients with paired samples, all four VOCs in the CRS panel and all three VOCs in the ICANS panel lost discriminative AUC for severe toxicity by Day +1 (CRS panel: 0.67–0.76 at Day −1 to 0.50–0.58 at Day +1; ICANS panel: 0.68–0.71 to 0.55–0.62; Supplementary Figure 5). For most panel VOCs, paired Wilcoxon signed-rank tests showed no significant change in absolute concentration between timepoints (CRS p = 0.113–0.692; ICANS p = 0.120–0.173), indicating that the AUC drop reflected reshuffling of patient ranks within the severe and non-severe groups rather than uniform concentration changes. Nonanal was the exception, falling significantly between timepoints (p = 0.022).

Three VOCs gained discriminative AUC at Day +1: β-pinene for severe ICANS (0.52 → 0.72, Δ +0.20), and 1,2,4-trimethylbenzene (0.59 → 0.66) and 2-heptanone (0.59 → 0.66) for severe CRS. Of these, only 2-heptanone showed a significant within-patient concentration increase (paired Wilcoxon p = 6 × 10⁻□; Supplementary Figure 6); β-pinene, 1,2,4-trimethylbenzene, α-pinene, and acetic acid did not (all p > 0.1). The Day +1 gains therefore reflect both absolute concentration shifts (2-heptanone) and rank reshuffling between severity groups (the remaining VOCs), consistent with rapid metabolic remodeling during CAR T-cell expansion.

### Day −1 VOC panel outperforms the published mEASIX score

mEASIX is a published clinical risk score (LDH × CRP / platelet count) that integrates pre-infusion endothelial-injury and inflammation biomarkers and has been validated as a predictor of grade ≥2 CRS after CAR T-cell therapy. We compared the day −1 CRS and ICANS panels with mEASIX in the same 26 patients (Figure 4). In our cohort, mEASIX achieved AUCs of 63.8% for grade ≥2 CRS and 50.6% for grade ≥2 ICANS. The 4-VOC CRS panel had numerically higher AUC than mEASIX (ΔAUC +18.7 percentage points, 95% CI −17.4 to +53.7; one-sided DeLong p = 0.15), without reaching statistical significance, likely reflecting the limited number of severe CRS events (n = 10). The 3-VOC ICANS panel outperformed mEASIX (ΔAUC +35.7 percentage points, 95% CI +6.4 to +65.0; one-sided DeLong p = 0.008). The VOC LOOCV score and log□(mEASIX) were essentially uncorrelated (Spearman ρ = 0.09 for CRS, p = 0.65; ρ = −0.07 for ICANS, p = 0.73), indicating that breath VOCs and the inflammation-endothelial composite capture distinct biology. A combined VOC + mEASIX SMAGS-LASSO LOOCV model did not improve on VOC alone (CRS 82.5% → 82.5%; ICANS 86.3% → 86.3%), consistent with the small severe-event count absorbing the additional parameter. The clinical implication is that breath VOCs identify pre-infusion risk that the routine endothelial-inflammation laboratory composite does not detect; whether the two combine usefully will be tested in the larger validation cohorts.

**Figure 4.**
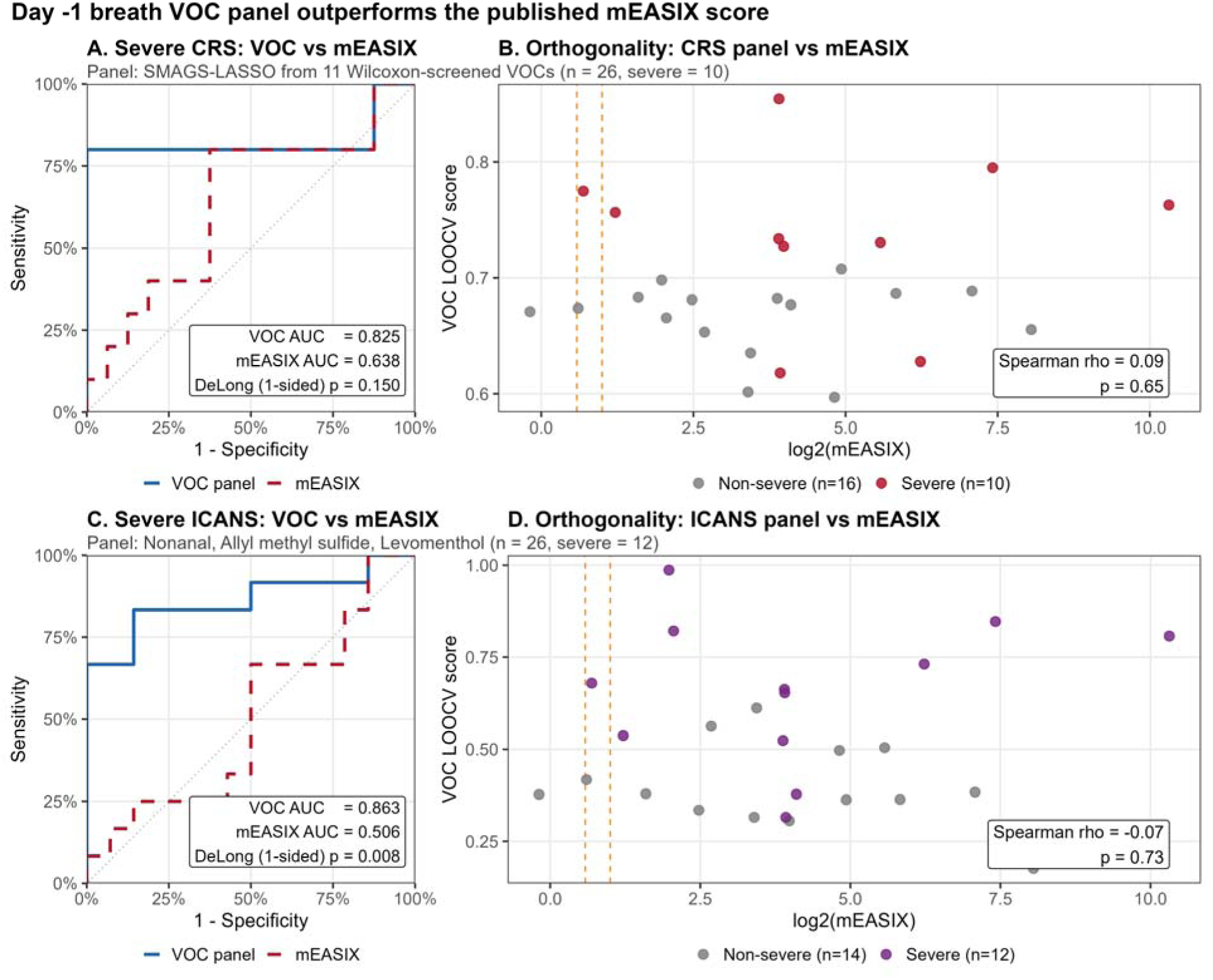
Day −1 SMAGS-LASSO VOC panel versus the published mEASIX score for grade ≥2 CRS and ICANS prediction. (A, C) LOOCV ROC curves comparing the VOC panel (blue/purple, solid), mEASIX as a continuous predictor (red, dashed), and a combined VOC + logD(mEASIX) SMAGS-LASSO LOOCV model (green, dotted). (B, D) Per-patient orthogonality: VOC LOOCV score versus logD(mEASIX) with grade ≥2 toxicity status overlaid. Vertical lines mark the published mEASIX risk thresholds (low/medium = logD4; medium/high = logD10).

## Discussion

In this proof-of-concept study, pre-infusion exhaled breath VOCs independently predicted severe grade ≥2 CRS and severe grade ≥2 ICANS in patients with r/r large B-cell lymphoma treated with axi-cel CAR T-cell therapy. A non-overlapping 4-VOC CRS panel and 3-VOC ICANS panel risk stratified patients to low-, intermediate- and high-risk tertiles, with 89% of high-risk group developing severe toxicity at faster onset than those in the low-risk group. Our pre-infusion model outperformed the published composite mEASIX score. These findings provide early evidence that non-invasive breath analysis can identify patients at substantially elevated risk for severe CAR T-cell toxicity before treatment delivery. If validated in larger prospective cohorts, breath analysis could enable preemptive intervention in high-risk patients.

Several panel VOCs are mechanistically linked to pathways related to lymphodepletion-induced oxidative stress and microbial perturbations in the peri-CAR T-cell therapy period. Aldehydes such as heptanal and nonanal arise from free radical-mediated peroxidation of polyunsaturated fatty acids,^16^ and benzaldehyde from lipid hydroperoxide-driven degradation of phenylalanine^17^ or microbial transamination of phenylalanine in the gut^18^. α- and β-pinenes reach exhaled breath through a composite of exogenous intake after hepatic biotransformation, gut microbial production^13^ and release from lipophilic stores. The day +1 emergence of pinenes in our cohort is consistent with well-documented gut microbiome perturbation that follows lymphodepletion^19^ or altered hepatic clearance during early CAR T-cell expansion.

Our findings extend prior plasma metabolomics studies of CAR T-cell toxicity to the breath compartment. In a cohort of B-cell lymphoma patients receiving CD19-directed CAR T-cell therapy, Jalota et al. reported that low pretreatment plasma proline, glycine, and isoursodeoxycholate were associated with severe CRS (grade≥ 2), and low hydroxyproline with higher-grade ICANS^11^ Isoursodeoxycholate is a secondary bile acid produced exclusively by gut bacterial 7α-dehydroxylation. This study also showed that plasma metabolites can predict earlier onset of toxicities. Although the specific metabolites differ, the VOCs we identified include benzaldehyde, which can form via lipid-hydroperoxide-driven phenylalanine degradation or microbial aminotransferase activity, and the monoterpenes β-pinene and α-pinene, with up to 40% of breath β-pinene variance attributable to gut microbiota composition^13^. Similar to Jalota et al., we also observed that pre-treatment VOCs predicted not only toxicity severity but also earlier onset, with Cox HR per 1-SD increment of 3.8 for severe CRS and 4.36 for severe ICANS. Whether these plasma and breath signatures reflect overlapping or independent biological events warrants direct comparison in larger cohorts.

Breath volatilome relevant to severe toxicity changes substantially during the peri-infusion window as shown by the non-overlap in VOC pattern between day −1 and day +1 samples. Per-VOC univariate AUCs shifted in opposite directions between the two timepoints: lipid peroxidation aldehydes such as heptanal and nonanal lost discriminative signal by day +1 while monoterpenes gained signal. The day +1 emergence of monoterpenes (β-pinene, α-pinene) and the short-chain fatty acid precursor acetic acid is consistent with early-phase post-lymphodepletion gut microbiome remodeling that starts within days of lymphodepletion and peaks around day +14^19,20^

## Limitations

This study has several limitations that should be acknowledged. First, the sample size was small (n = 26 patients; 10 severe CRS events and 12 severe ICANS events), and external validation in larger, independent cohorts will be required before clinical application. LASSO regularization, leave-one-out cross-validation, and Harrell bootstrap optimism correction partially mitigate the small-sample concern. Second, the breath analysis was targeted to 40 VOCs, and an untargeted analytical approach with broader compound coverage may identify additional candidate biomarkers. Third, breath VOC composition is influenced by diet, medications, and comorbidities; although we applied ambient-air subtraction and a 24-hour dietary recall, residual confounding from environmental and behavioral sources cannot be excluded. Finally, our findings derive from a single institution and a homogeneous patient population, and generalizability to other CAR T-cell products, disease contexts, and treatment centers will require multi-institutional prospective validation.

## Conclusion

Exhaled breath VOC analysis identifies biologically coherent, non-invasive and syndrome specific predictors of grade ≥2 CAR T-cell therapy toxicities across two distinct peri-infusion windows. If validated in prospective multi-center validation, breath analysis could enable pre-infusion risk stratification and identify patients who would benefit from preemptive intervention for severe CRS and ICANS.

## Data Availability

All data produced in the present work are contained in the manuscript

## References

1. Neelapu SS, Tummala S, Kebriaei P, et al. Chimeric antigen receptor T-cell therapy — assessment and management of toxicities. Nat Rev Clin Oncol. 2018;15(1):47–62. doi:10.1038/nrclinonc.2017.148

2. Neelapu SS. Managing the toxicities of CAR T-cell therapy. Hematol Oncol. 2019;37 Suppl 1:48–52. doi:10.1002/hon.2595

3. Locke FL, Ghobadi A, Jacobson CA, et al. Long-term safety and activity of axicabtagene ciloleucel in refractory large B-cell lymphoma (ZUMA-1): a single-arm, multicentre, phase 1–2 trial. The Lancet Oncology. 2019;20(1):31–42. doi:10.1016/S1470-2045(18)30864-7

4. Lee DW, Santomasso BD, Locke FL, et al. ASTCT Consensus Grading for Cytokine Release Syndrome and Neurologic Toxicity Associated with Immune Effector Cells. Biology of Blood and Marrow Transplantation. 2019;25(4):625–638. doi:10.1016/j.bbmt.2018.12.758

5. Brudno JN, Kochenderfer JN. Advances in the mechanisms and management of CAR T-cell toxicities. Nat Rev Clin Oncol. 2024;21(7):501–521. doi:10.1038/s41571-024-00903-0

6. Teachey DT, Bishop MR, Maloney DG, Grupp SA. Toxicity management after chimeric antigen receptor T cell therapy: one size does not fit “ALL.” Nat Rev Clin Oncol. 2018;15(4):218–218. doi:10.1038/nrclinonc.2018.19

7. Pennisi M, Sanchez-Escamilla M, Flynn JR, et al. Modified EASIX predicts severe cytokine release syndrome and neurotoxicity after chimeric antigen receptor T cells. Blood Adv. 2021;5(17):3397–3406. doi:10.1182/bloodadvances.2020003885

8. de Boer JW, Keijzer K, Pennings ERA, et al. Population-Based External Validation of the EASIX Scores to Predict CAR T-Cell-Related Toxicities. Cancers (Basel). 2023;15(22):5443. doi:10.3390/cancers15225443

9. Chou H, Godbeer L, Allsworth M, Boyle B, Ball ML. Progress and challenges of developing volatile metabolites from exhaled breath as a biomarker platform. Metabolomics. 2024;20(4):72. doi:10.1007/s11306-024-02142-x

10. Berna AZ, Akaho EH, Harris RM, et al. Reproducible Breath Metabolite Changes in Children with SARS-CoV-2 Infection. ACS Infect Dis. 2021;7(9):2596–2603. doi:10.1021/acsinfecdis.1c00248

11. Jalota A, Hershberger CE, Patel MS, et al. Host metabolome predicts the severity and onset of acute toxicities induced by CAR T-cell therapy. Blood Advances. 2023;7(17):4690–4700. doi:10.1182/bloodadvances.2022007456

12. Berna AZ, DeBosch B, Stoll J, Odom John AR. Breath Collection from Children for Disease Biomarker Discovery. J Vis Exp. 2019;(144). doi:10.3791/59217

13. The gut microbiota shapes the human and murine breath volatilome: Cell Metabolism. Accessed April 30, 2026. https://www.cell.com/cell-metabolism/fulltext/S1550-4131(25)00544-3

14. Ghasemi SM, Gu C, Fahrmann JF, et al. A Novel Sensitivity Maximization at a Given Specificity Method for Binary Classifications. Cancer Prev Res (Phila). 2025;18(3):117–123. doi:10.1158/1940-6207.CAPR-24-0236

15. SMAGS-LASSO: A Novel Feature Selection Method for Sensitivity Maximization in Early Cancer Detection | Cancer Epidemiology, Biomarkers & Prevention | American Association for Cancer Research. Accessed May 1, 2026. https://aacrjournals.org/cebp/article/34/12/2259/767337/SMAGS-LASSO-A-Novel-Feature-Selection-Method-for?guestAccessKey=

16. Esterbauer H, Schaur RJ, Zollner H. Chemistry and biochemistry of 4-hydroxynonenal, malonaldehyde and related aldehydes. Free Radic Biol Med. 1991;11(1):81–128. doi:10.1016/0891-5849(91)90192-6

17. Hidalgo FJ, Zamora R. Formation of phenylacetic acid and benzaldehyde by degradation of phenylalanine in the presence of lipid hydroperoxides: New routes in the amino acid degradation pathways initiated by lipid oxidation products. Food Chem X. 2019;2:100037. doi:10.1016/j.fochx.2019.100037

18. Nierop Groot MN null, de Bont JAM null. Conversion of phenylalanine to benzaldehyde initiated by an aminotransferase in lactobacillus plantarum. Appl Environ Microbiol. 1998;64(8):3009-3013. doi:10.1128/AEM.64.8.3009-3013.1998

19. Schluter J, Peled JU, Taylor BP, et al. The gut microbiota is associated with immune cell dynamics in humans. Nature. 2020;588(7837):303–307. doi:10.1038/s41586-020-2971-8

20. Smith M, Dai A, Ghilardi G, et al. Gut microbiome correlates of response and toxicity following anti-CD19 CAR T cell therapy. Nat Med. 2022;28(4):713–723. doi:10.1038/s41591-022-01702-9

