## Supplement for "Pre-infusion Exhaled breath volatile organic compounds predict severe CRS and ICANS after CAR T-cell therapy"

**Supplementary Materials**

**Supplementary Figure 1. CAR T-cell toxicity timeline**

Swimmer plot showing CRS and ICANS grade trajectory for each patient with breath sampling timepoints overlaid.

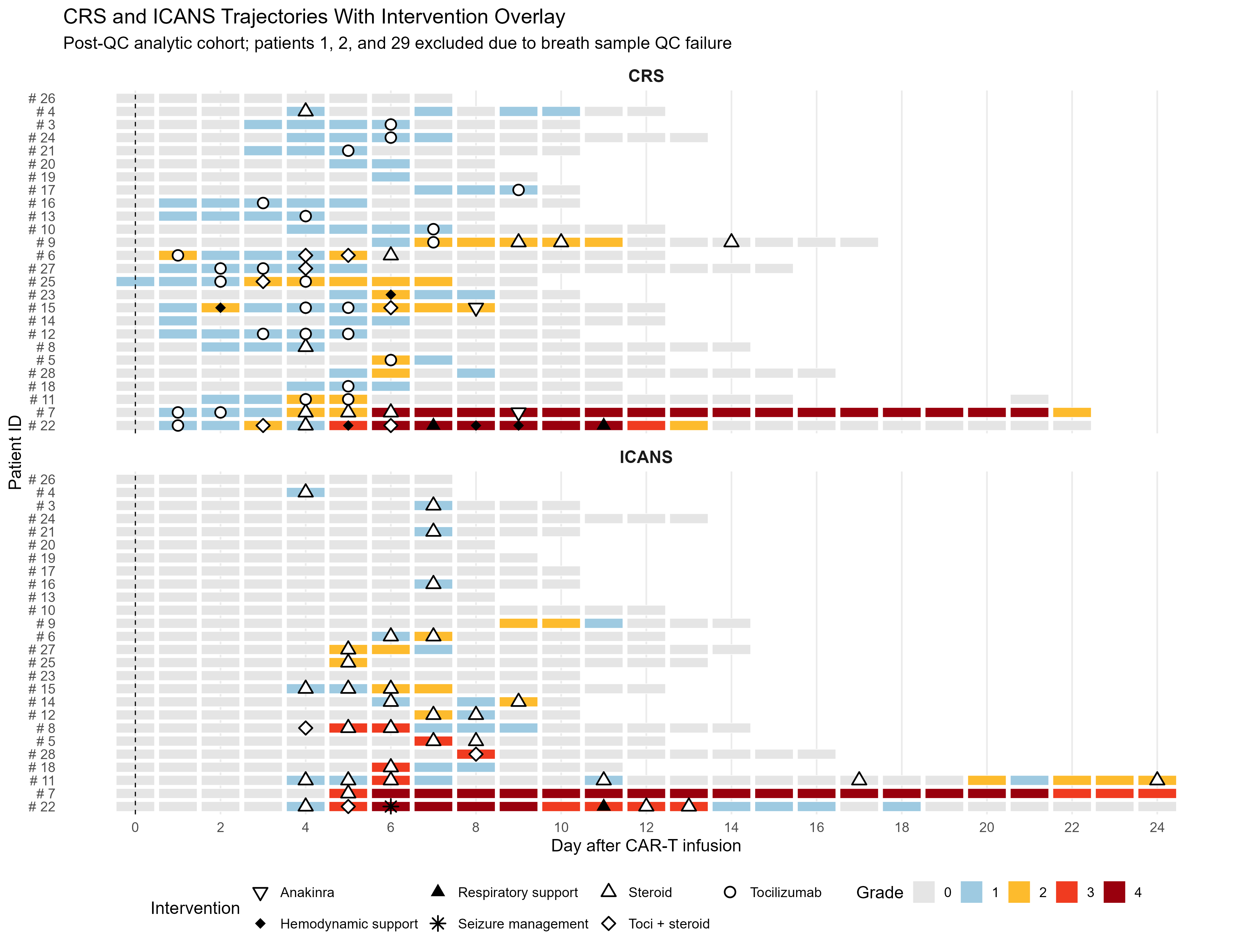

**Supplementary Figure 2. Bootstrap stability of day −1 panels**

Selection frequency and coefficient distribution for each panel VOC across 1,000 bootstrap iterations.

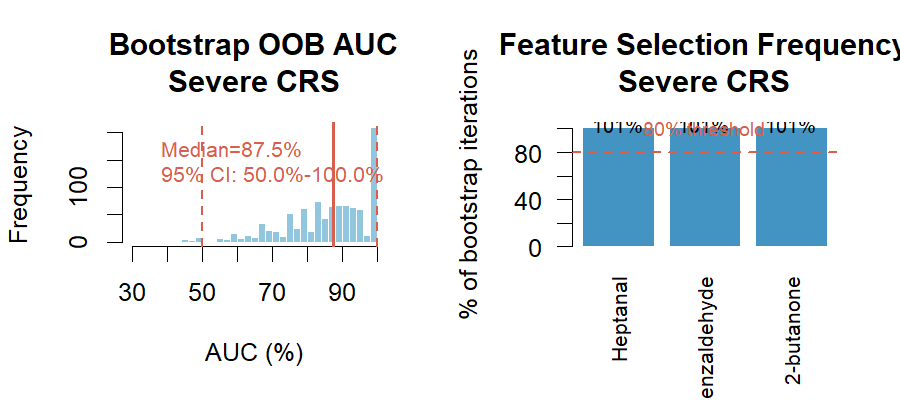

**A.** CRS panel bootstrap stability.

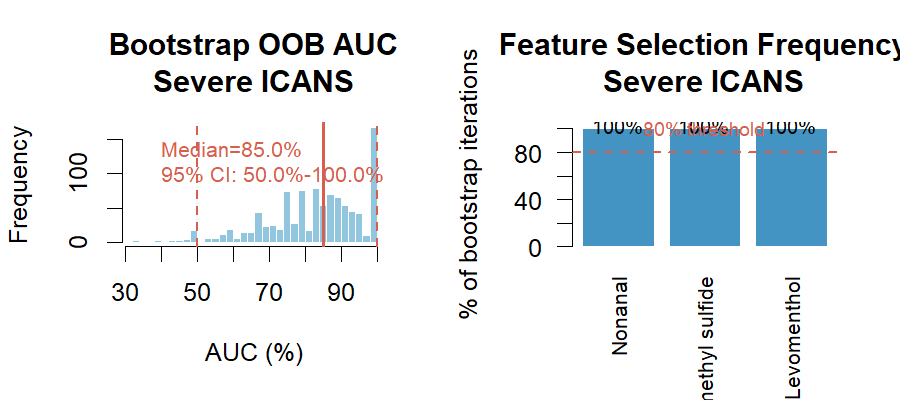

**B.** ICANS panel bootstrap stability.

**Supplementary Figure 3. Permutation tests for day −1 panels**

Distribution of LOOCV AUCs from 1,000 label permutations.

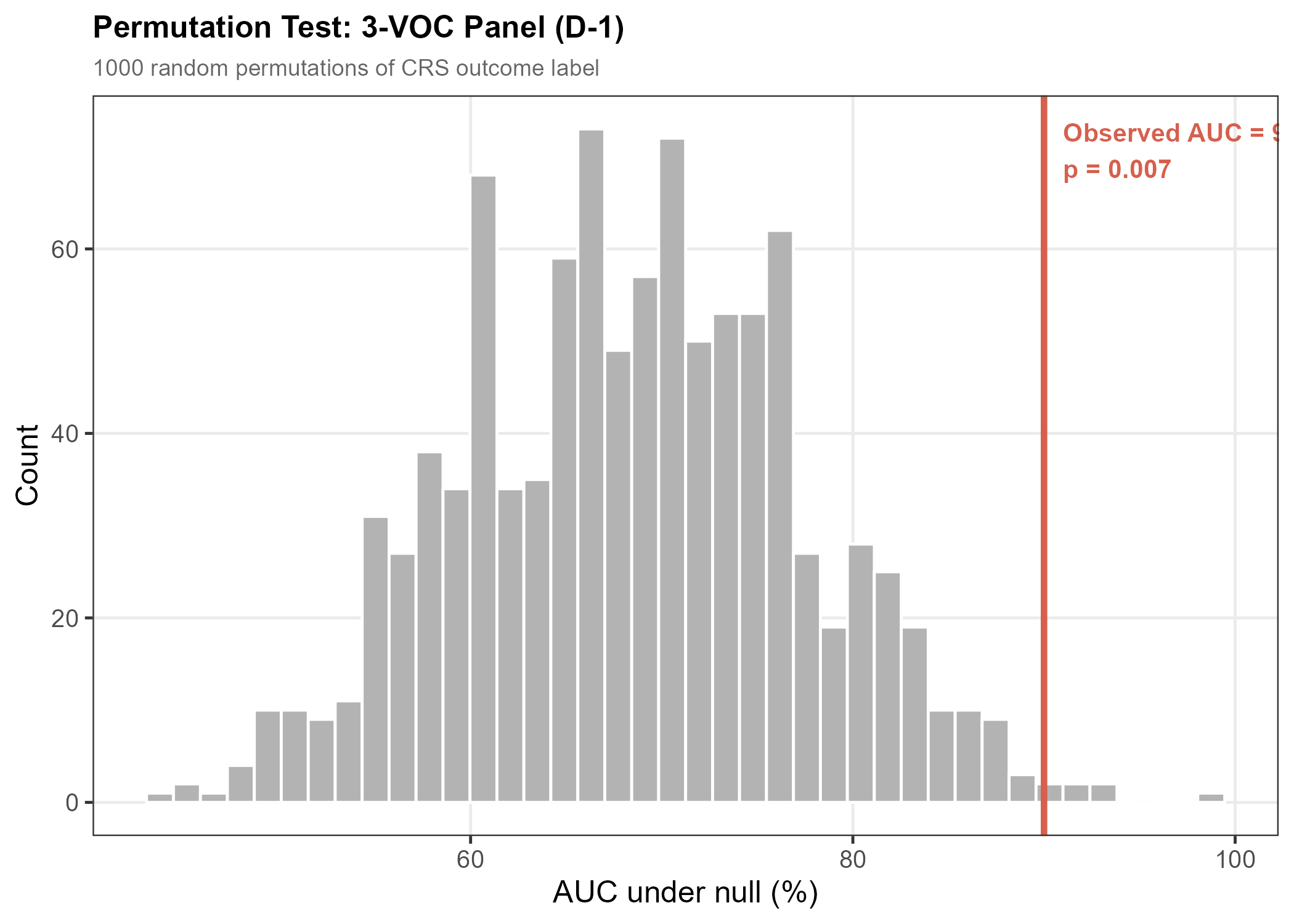

**A.** CRS panel permutation test (observed AUC 82.5%, p=0.007).

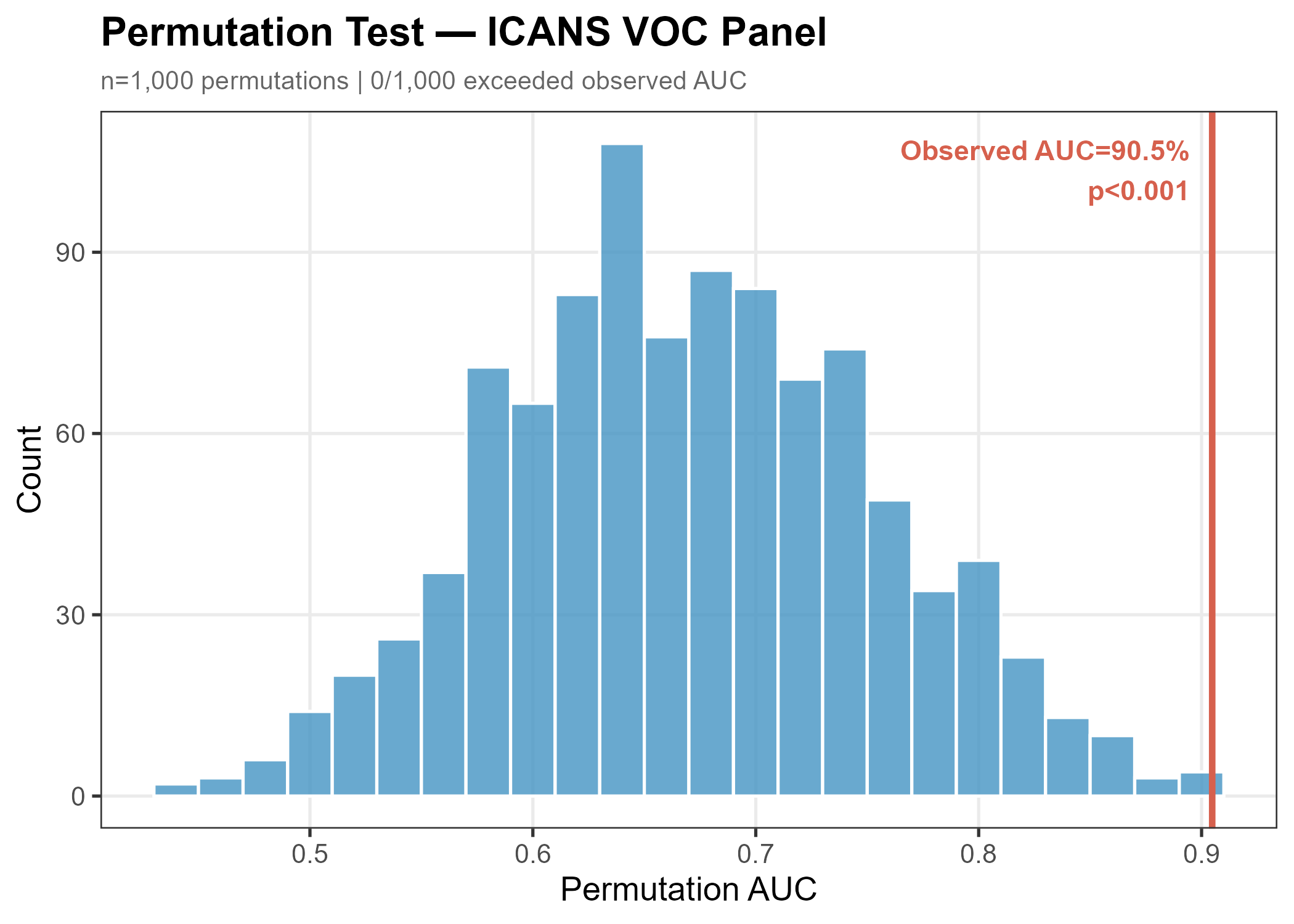

**B.** ICANS panel permutation test (observed AUC 86.3%, p<0.001).

**Supplementary Figure 4. VOC heatmaps**

Per-patient z-scores of all candidate VOCs (AUC ≥0.65 in screening), clustered by sample and compound.

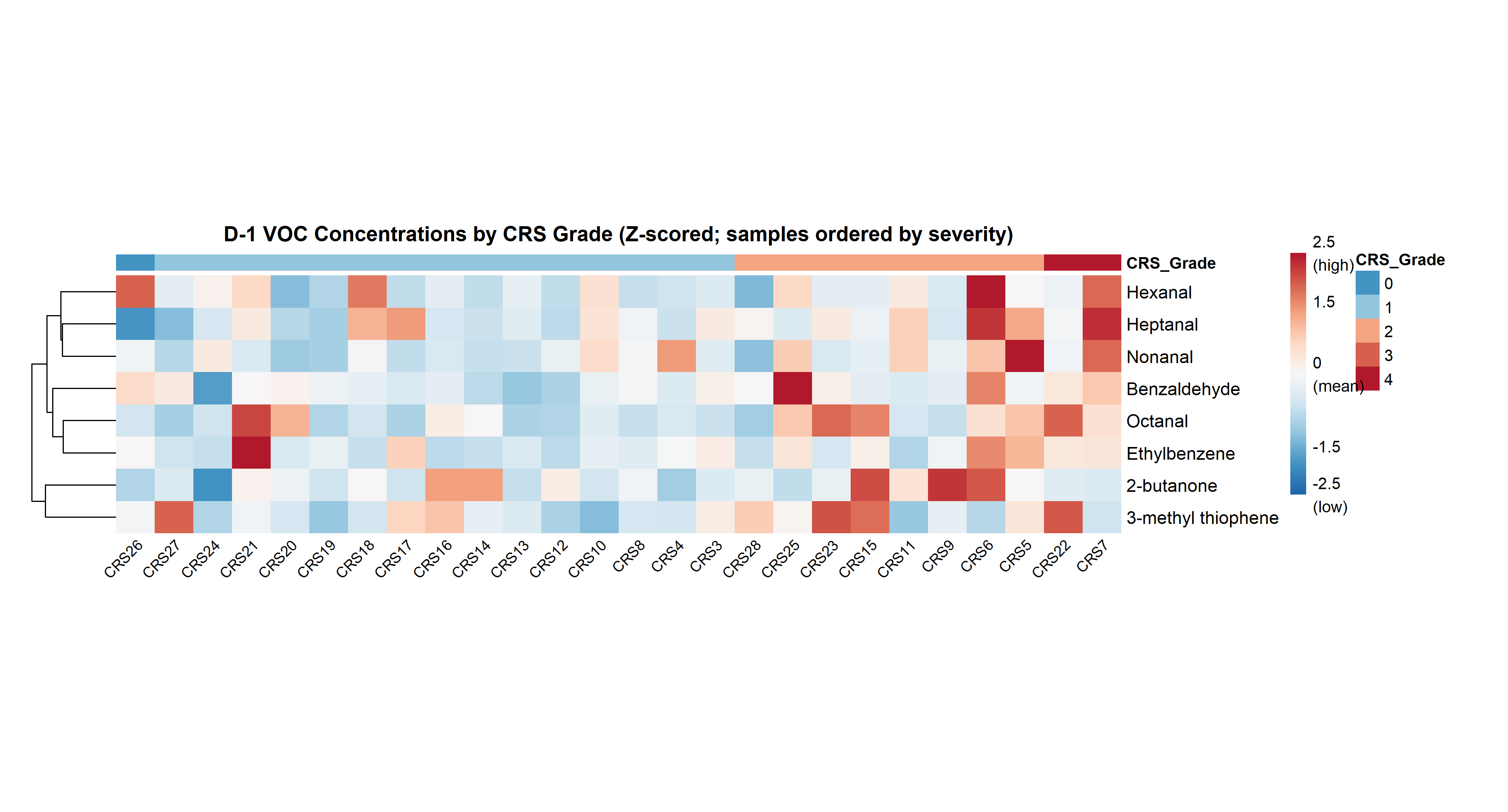

**A.** CRS candidate VOCs.

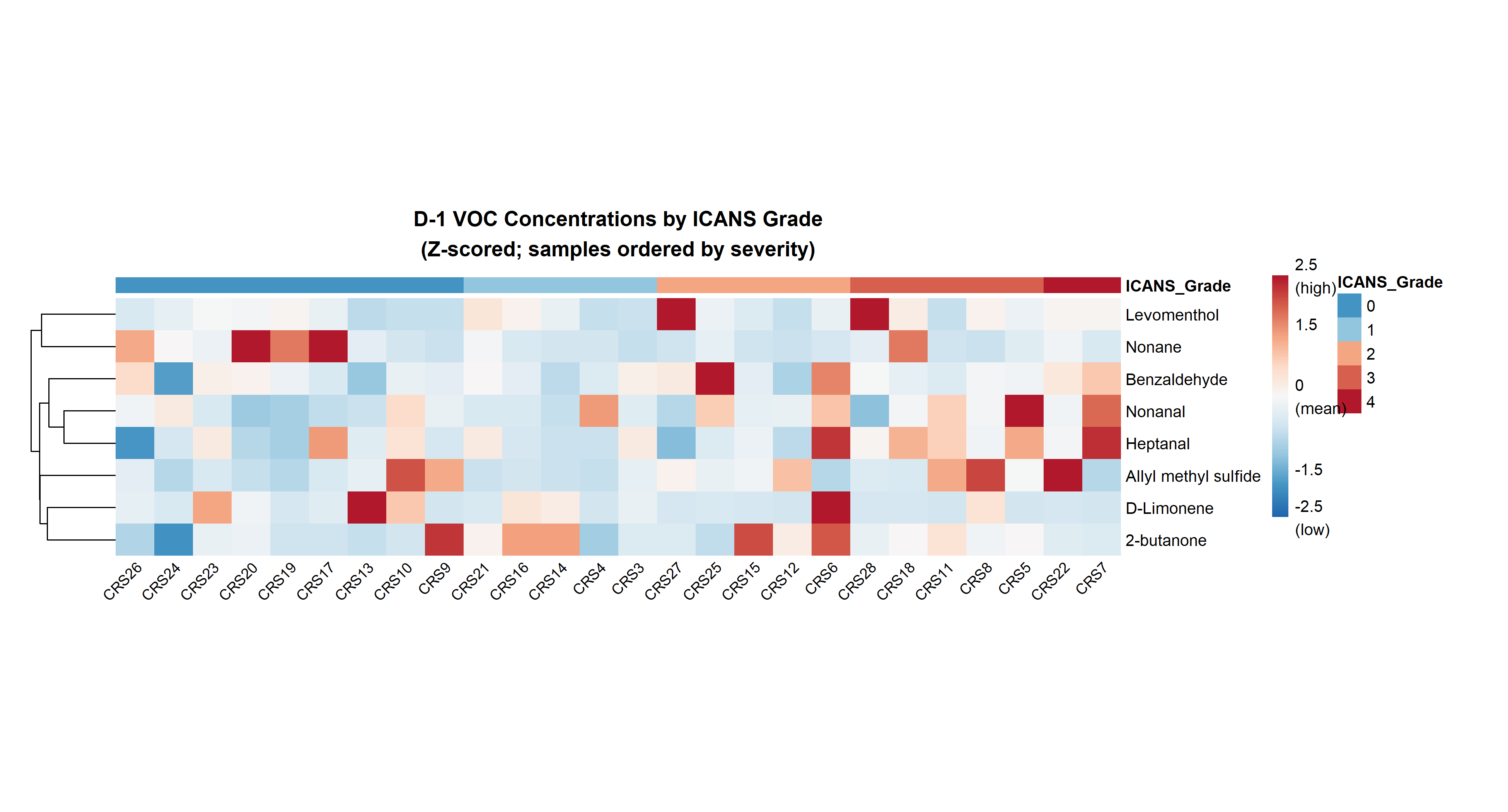

**B.** ICANS candidate VOCs.

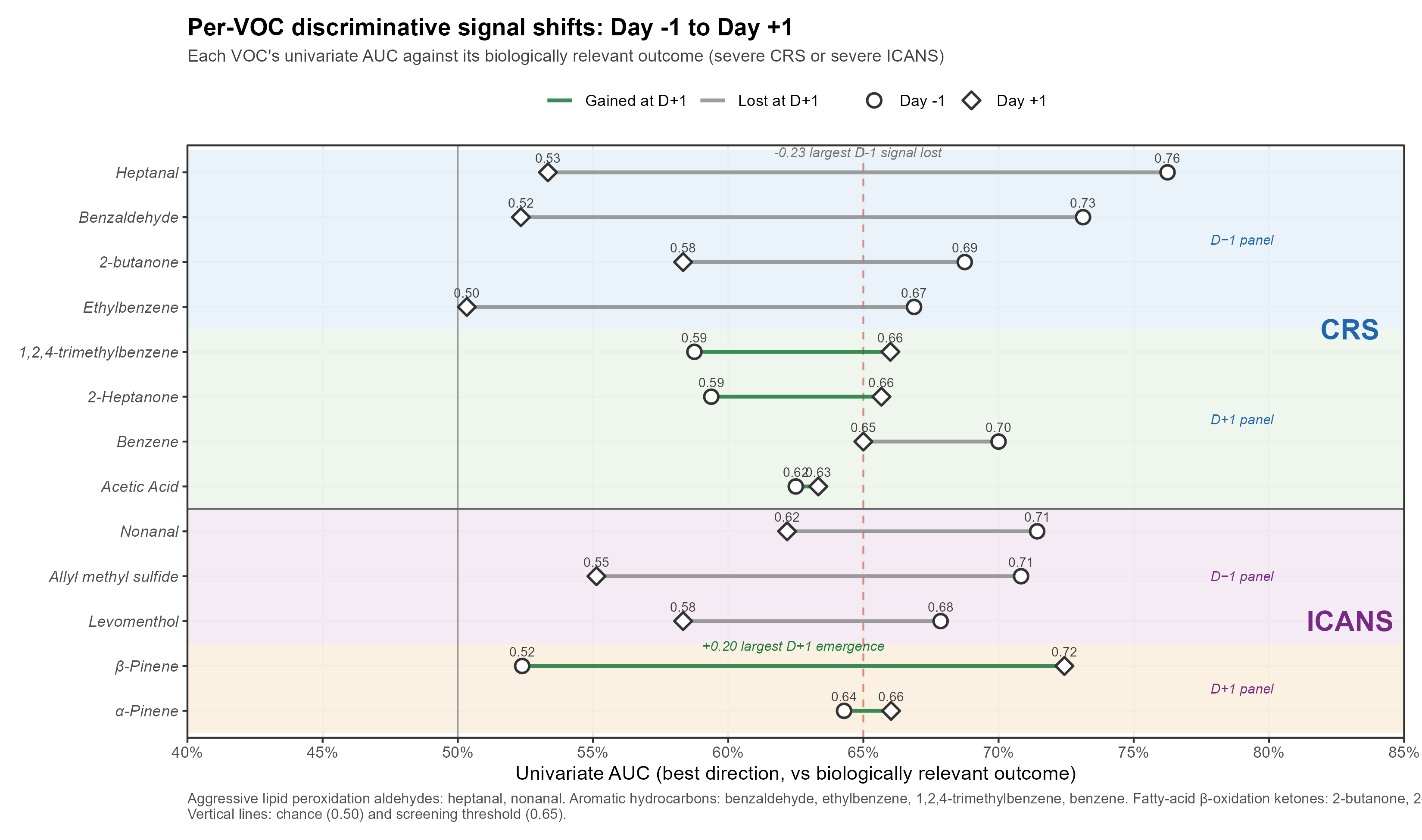

**Supplementary Figure 5.** Per-VOC AUC shift between day −1 and day +1. Each panel VOC is shown as a circle (day −1 AUC) connected by a line to a diamond (day +1 AUC). Lines are colored by biological pathway theme when the signal strengthens at day +1 (terpene/short-chain fatty acid emergence) and grey when the signal weakens (aldehyde/oxidative-stress signal loss). Reference lines mark AUC=0.50 (chance) and 0.65 (screening threshold).

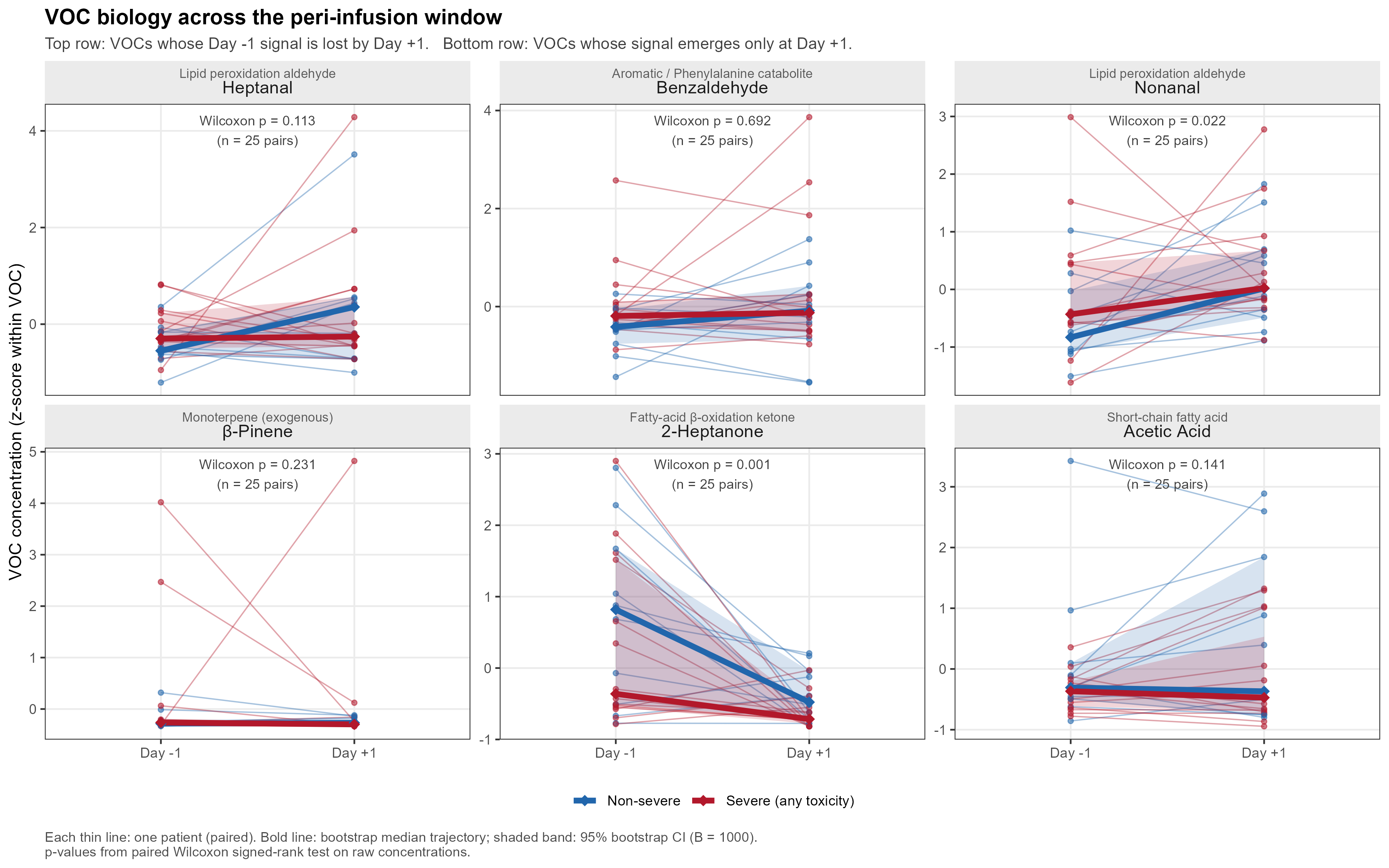

**Supplementary Figure 6.** Volatile organic compound (VOC) biology across the peri-infusion window. Trajectories of six representative panel VOCs from the day before lymphodepletion (day −1) to the day after CAR T-cell infusion (day +1) in 25 patients with paired samples. Top row shows VOCs whose Day −1 univariate signal for severe toxicity is lost by day +1: heptanal and nonanal, and benzaldehyde. Bottom row shows VOCs whose signal emerges only at Day +1: β-pinene, 2-heptanone, and acetic acid. For visual comparability, each VOC's concentrations are z-scored within VOC across all paired observations. Thin lines represent individual patients (paired Day −1 to Day +1); bold lines show the bootstrap median trajectory for non-severe (blue) and severe-any-toxicity (red) groups, and shaded ribbons show the 95% bootstrap confidence interval of the median (B = 1000 resamples). p-values are from paired Wilcoxon signed-rank tests on raw (un-z-scored) concentrations.

**Supplementary Table 1. Candidate VOC AUCs**

All candidate VOCs (AUC ≥0.65) for grade ≥2 CRS and grade ≥2 ICANS at day −1, with Wilcoxon p-values and Benjamini–Hochberg adjusted q-values. Available as a separate data file (VOC_D_1_CRS_Wilcoxon_AUC.csv, VOC_D_1_ICANS_Wilcoxon_AUC.csv).

**Supplementary Note 1. Global breath volatilome shift D−1 → D+1**

To complement the targeted predictive analyses, we examined the global paired shift in the breath volatilome between day −1 and day +1 in the 24 patients with paired samples available at both timepoints (one day +1 sample missing; one day −1 PCA outlier excluded by an a priori |z(PC1)|>3 rule). The volatilome shifted significantly between timepoints (PERMANOVA on Euclidean distance, blocked by patient: p<0.001, R²=0.042), but the magnitude of the per-patient shift did not differ between patients who developed grade ≥2 toxicity and those who did not (Mann–Whitney p=0.65). Three VOCs crossed Benjamini–Hochberg q<0.05 in the per-VOC paired Wilcoxon analysis: 2-Heptanone (↓, log₂FC −1.91), Styrene (↑, log₂FC +0.28), and 1-Hexanol, 2-ethyl- (↑, log₂FC +0.34). The CAR-T effect on the global volatilome is therefore small (R² ≈ 4%) and concentrated in a handful of compounds rather than a coordinated metabolome-wide rearrangement; this is consistent with the predictive analyses, which show that distinct subsets of VOCs carry the toxicity signal at the two timepoints.

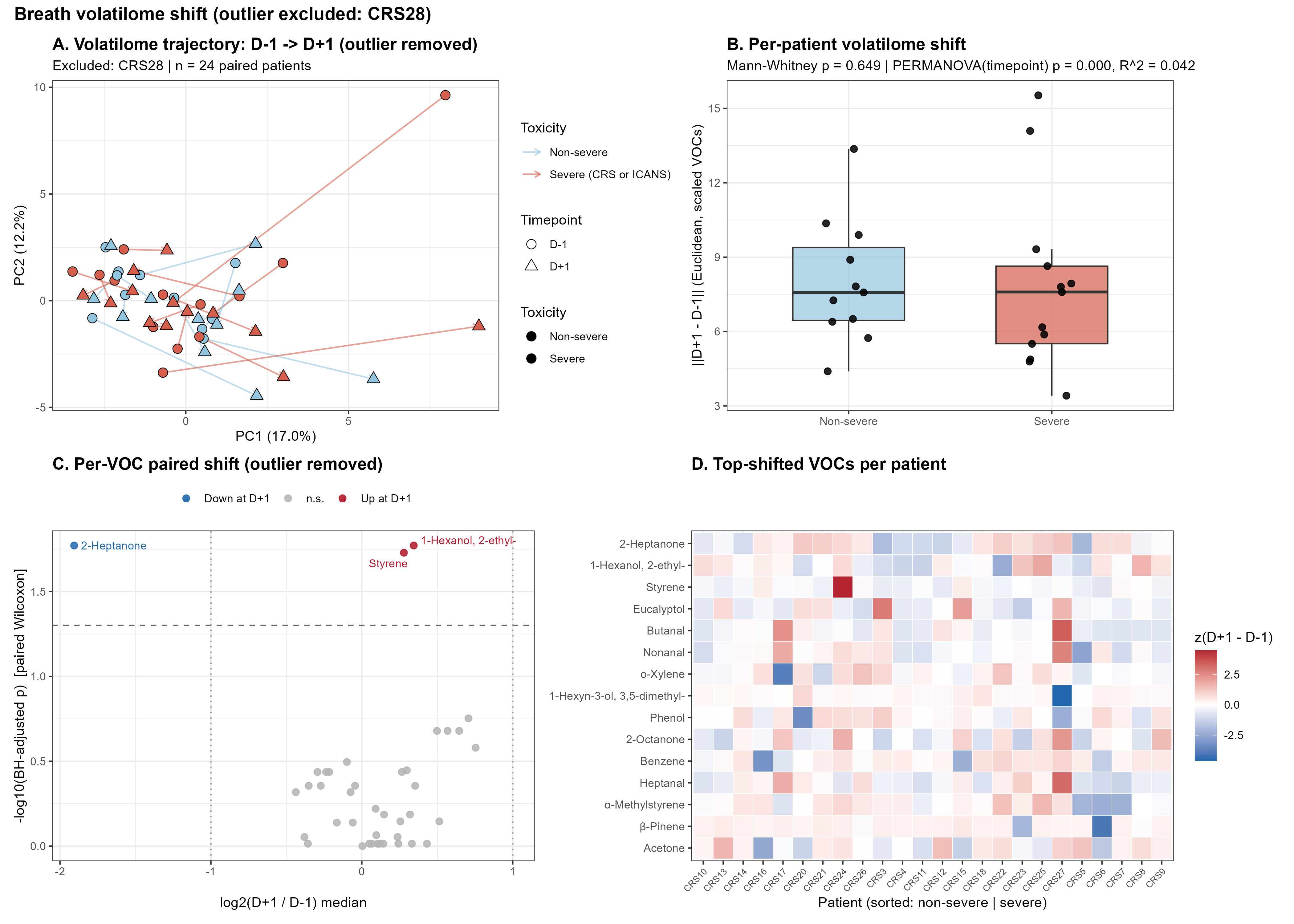

**Supplementary Figure 7.** Paired day −1 → day +1 volatilome shift (n=24 paired patients; CRS28 excluded as PCA outlier). (A) PCA biplot of standardized VOC profiles with arrows connecting each patient's day −1 (circle) and day +1 (triangle) samples; color = severe toxicity status. (B) Per-patient Euclidean shift magnitude in the scaled VOC space, stratified by severe-toxicity status. (C) Volcano plot of the per-VOC paired Wilcoxon test (log₂(D+1/D−1) median vs −log₁₀ BH-adjusted p). (D) Heatmap of the 15 most-shifted VOCs per patient (z-scored Δ = D+1 − D−1); patients ordered non-severe → severe.

**Supplementary Figure 8. Bootstrap optimism correction and 95% CI**

Harrell bootstrap optimism correction (1,000 iterations) for the day −1 SMAGS-LASSO panels. Each iteration: (i) draw a bootstrap sample of size n=26 with replacement; (ii) fit SMAGS-LASSO on the bootstrap sample; (iii) score both the bootstrap sample (apparent) and the original sample (test); (iv) optimism = apparent − test. The optimism-corrected AUC is the full-sample apparent AUC minus the mean optimism across 1,000 iterations. The 95% CI is the percentile CI of the bootstrap test AUC distribution. Both panels are robust: CRS optimism-corrected AUC 0.877 (95% CI 0.73–0.96) and ICANS 0.867 (95% CI 0.74–0.91); both lower bounds exceed the 0.65 screening threshold, and the bootstrap-corrected AUCs lie between the apparent (in-sample) and LOOCV AUCs as expected.

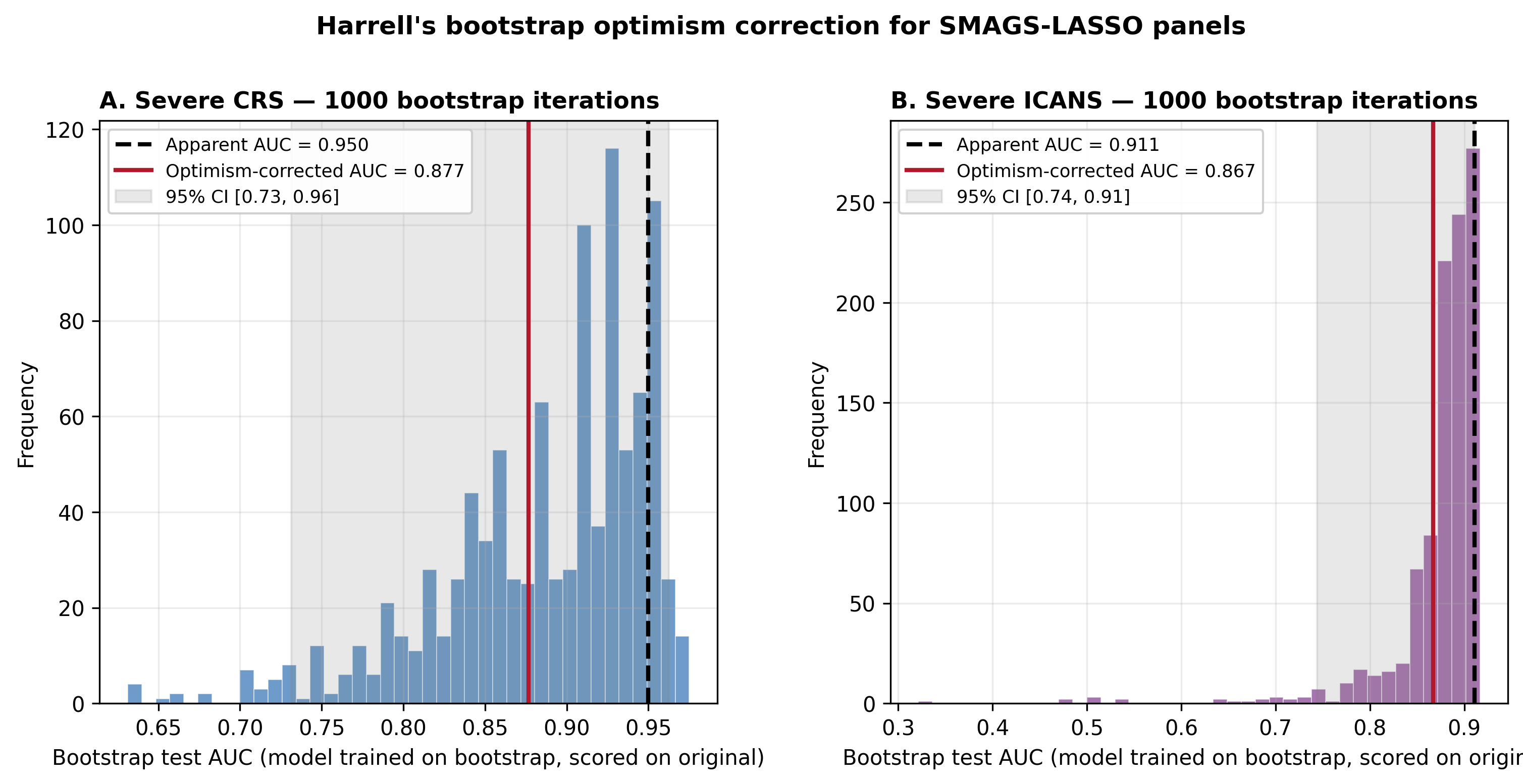

**Supplementary Figure 9.** Harrell bootstrap optimism correction (1,000 iterations) for the day −1 SMAGS-LASSO panels. (A) Severe CRS, 4-VOC panel: apparent AUC 0.950 (dashed black), optimism-corrected AUC 0.877 (red), 95% CI [0.73, 0.96] (grey shading). (B) Severe ICANS, 3-VOC panel: apparent AUC 0.911, optimism-corrected AUC 0.867, 95% CI [0.74, 0.91]. Histograms show the distribution of bootstrap test AUCs (model trained on bootstrap, scored on the original cohort) across 1,000 iterations.
